# BEGA-UNet: Boundary-Explicit Guided Attention U-Net with Multi-Scale Feature Aggregation for Colonoscopic Polyp Segmentation

**DOI:** 10.64898/2026.03.04.26347608

**Authors:** Tao Tong, Wen Zhang, Wanni Zu

## Abstract

Accurate polyp segmentation from colonoscopy images is critical for colorectal cancer prevention, yet the generalization of deep learning models under domain shift remains insufficiently explored. We propose Boundary-Explicit Guided Attention U-Net (BEGA-UNet), a boundary-aware segmentation architecture that introduces explicit edge modeling as a structural inductive bias to enhance both segmentation accuracy and cross-domain robustness. The framework integrates three components: an Edge-Guided Module (EGM) with learnable Sobel-initialized operators to capture boundary cues, a Dual-Path Attention (DPA) module that processes channel and spatial attention in parallel, and a Multi-Scale Feature Aggregation (MSFA) module to encode contextual information across multiple receptive fields. Evaluated on the combined Kvasir-SEG and CVC-ClinicDB benchmarks, BEGA-UNet achieves 88.53% Dice and 82.51% IoU, outperforming representative convolutional and transformer-based baselines. More importantly, cross-dataset evaluation demonstrates strong robustness under domain shift, with BEGA-UNet retaining 83.2% of its in-distribution performance—substantially higher than U-Net (64.5%), Attention U-Net (47.5%), and TransUNet (53.1%). In a zero-shot setting on an entirely unseen dataset, the model further maintains 72.6% performance retention. Comprehensive ablation studies indicate that explicit boundary modeling plays a central role in improving generalization, while multi-scale context aggregation further stabilizes performance across domains. Feature distribution analyses support this observation by showing that edge-oriented representations exhibit markedly reduced cross-domain variability compared to appearance-driven features. Overall, BEGA-UNet provides an effective and interpretable solution for robust polyp segmentation, demonstrating that explicit boundary modeling serves as a critical inductive bias for ensuring reliability under clinical domain shifts.

## 1. Introduction

Colorectal cancer (CRC) represents the third most commonly diagnosed malignancy and the second leading cause of cancer-related mortality worldwide, accounting for approximately 1.9 million new cases and 935,000 deaths annually ^[1]^. The adenoma-carcinoma sequence provides a critical window for early intervention, as benign colorectal polyps progressively transform into malignant tumors over 10-15 years ^[2]^. Colonoscopy remains the gold standard for polyp detection and removal, with polypectomy reducing CRC mortality by up to 53% ^[3]^. However, the adenoma miss rate during routine colonoscopy ranges from 6% to 27%, primarily due to operator fatigue, suboptimal bowel preparation, and difficulty detecting flat or diminutive polyps ^[4[5.^

Computer-aided detection and diagnosis (CADe/CADx) systems have emerged as promising tools to enhance colonoscopic examination quality ^[6]^, with recent cross-domain polyp segmentation networks leveraging boundary-focused distillation and large vision models to improve robustness under distribution shift ^[7]^. Deep learning approaches have revolutionized medical image segmentation, with recent theoretical syntheses highlighting rapid advances in architectural inductive biases, generalization strategies, and robustness modeling ^[8]^.

The seminal U-Net architecture ^[9]^, featuring symmetric encoder-decoder pathways with skip connections, has become the cornerstone of biomedical image segmentation. Subsequent innovations have enhanced U-Net through various strategies: Attention U-Net ^[10]^ introduces attention gates; U-Net++ ^[11]^ employs nested dense skip pathways; PraNet ^[12]^ proposes parallel reverse attention; and TransUNet ^[13]^ integrates Transformer encoders for long-range dependencies.

Despite these advances, accurate polyp segmentation remains challenging due to three inherent difficulties. **(1) Ambiguous boundaries:** Polyp margins often exhibit gradual intensity transitions that blend with adjacent healthy mucosa, particularly for sessile and flat polyps ^[14]^. **(2) Appearance heterogeneity:** Polyps demonstrate substantial variability in size (diminutive <5mm to large >20mm), shape, color, and texture across patients and anatomical locations ^[15]^. **(3) Imaging artifacts:** Colonoscopy images frequently contain specular reflections, motion blur, and residual fecal matter, creating complex backgrounds ^[16]^.

Current approaches exhibit three methodological limitations: (1) reliance on implicit boundary learning, where standard convolutions may inadequately capture fine-grained edge information critical for precise delineation ^[17]^; (2) sequential attention designs that risk creating information bottlenecks and attenuating boundary signals; and (3) single-scale feature representations insufficient for characterizing the substantial size variability of polyps ^[18]^.

To address these limitations, this work proposes **BEGA-UNet** (Boundary-Explicit Guided Attention U-Net), a novel architecture explicitly leveraging boundary information and multi-scale contextual cues.

### Our principal contributions are threefold

First, we present BEGA-UNet, a unified framework where explicit boundary modeling serves as a structural prior to guide attention-based segmentation; the Edge-Guided Module (EGM) combines Sobel-initialized operators with end-to-end learning, bridging classical edge detection and deep feature learning. Second, we provide systematic empirical evidence that explicit boundary features exhibit stronger domain invariance than implicit learned representations; cross-dataset experiments demonstrate that BEGA-UNet maintains 83.2% of in-distribution performance under domain shift, compared to 47.5–64.5% for established alternatives. Third, we introduce dual-protocol ablation analysis that reveals functional subsumption among modules, wherein explicit boundary modeling subsumes attention-based boundary preservation while boundary and multi-scale modules exhibit the highest complementarity (62.3% additivity retention); these interaction patterns provide transferable design principles for boundary-aware architectures.

## 2. Related Work

### 2.1 Deep Learning for Polyp Segmentation

Fully convolutional networks (FCNs) ^[19]^ marked a paradigm shift in semantic segmentation. U-Net ^[9]^ extended this with encoder-decoder architecture and skip connections, establishing the foundation for medical image segmentation. Subsequent advances include attention-based methods (PraNet ^[12]^, SANet ^[20]^, UACANet ^[23]^), efficient architectures (HarDNet-MSEG ^[21]^), and Transformer integration (Polyp-PVT ^[22]^, SSFormer ^[24]^). M^2^SNet ^[46]^ employs multi-scale subtraction for implicit boundary enhancement. While these methods achieve competitive benchmark performance, marginal in-distribution gains motivate our focus on cross-domain generalization as the primary evaluation criterion.

### 2.2 Attention Mechanisms

Attention mechanisms enhance feature representations effectively and have been increasingly integrated into encoder-decoder architectures for medical image segmentation, demonstrating improved feature selectivity across diverse clinical tasks ^[25]^. SE-Net ^[26]^ introduces channel attention through squeeze-and-excitation. Convolutional Block Attention Module (CBAM) ^[27]^ combines channel and spatial attention sequentially. DANet ^[28]^ employs dual attention for channel and position dependencies. In polyp segmentation, ACSNet ^[29]^ proposes adaptive context selection, while CaraNet ^[30]^ introduces channel-wise feature recalibration. Most existing methods process attention types sequentially, potentially creating information bottlenecks. This sequential design limits the interaction between different attention types and may cause gradient degradation in deep networks.

### 2.3 Boundary-Aware Segmentation

Explicit boundary modeling improves segmentation accuracy for objects with indistinct edges. Recent boundary-aware segmentation frameworks have explored edge-enhanced feature learning to refine structural delineation under ambiguous edge conditions ^[31]^. BASNet ^[32]^ incorporates boundary-aware refinement. In polyp segmentation, boundary-aware approaches adopt divergent strategies. EU-Net ^[33]^ employs fixed Canny operators for edge detection, providing explicit but non-adaptive boundary cues that cannot adjust to dataset-specific characteristics. M^2^SNet ^[46]^ enhances boundaries through multi-scale subtraction, where boundary signals emerge as byproducts of feature differencing rather than from dedicated edge operators. Multi-scale boundary-aware strategies have also been explored for capturing structural details across varying object scales ^[34]^. Our approach bridges the gap between fixed and implicit boundary strategies: EGM combines explicit edge modeling (via Sobel-initialized operators) with end-to-end learnability. Unlike implicit enhancement, this design forces the network to decouple structural boundary information from domain-specific appearance, fostering better generalization.

### 2.4 Multi-Scale Feature Learning

Multi-scale representations are essential for variable-size object segmentation. DeepLab series ^[35[36^ popularized atrous spatial pyramid pooling (ASPP). PSPNet ^[37]^ employs pyramid pooling for global context. Recent multi-scale architectures integrate convolutional attention with frequency-enhanced transformer encoders to capture hierarchical contextual dependencies across varying receptive fields ^[38]^. These approaches demonstrate that explicit multi-scale modeling benefits polyp segmentation significantly. Collectively, these advances primarily target in-distribution benchmark optimization. In contrast, this work emphasizes cross-dataset generalization as the primary evaluation criterion, hypothesizing that explicit boundary modeling provides domain-invariant priors with superior transferability.

## 3. Methodology

### 3.1 Architecture Overview

Design Philosophy. BEGA-UNet is guided by a central principle: explicit boundary modeling as a structural prior to constrain dense attention-based segmentation. This philosophy manifests through three interconnected mechanisms: (1) EGM extracts learnable boundary features serving as structural priors that provide explicit geometric constraints; (2) DPA employs parallel attention pathways to refine features while preserving boundary signal integrity, preventing over-attenuation inherent in sequential designs; and (3) MSFA propagates boundary constraints across multiple receptive field scales, ensuring effective guidance regardless of polyp size.

Figure 1 illustrates the overall architecture of BEGA-UNet, following an encoder-decoder paradigm with four encoder stages, an MSFA-enhanced bottleneck, and four symmetric decoder stages. EGM modules are integrated at each encoder stage, while DPA modules are deployed throughout both pathways.

**Figure 1.**
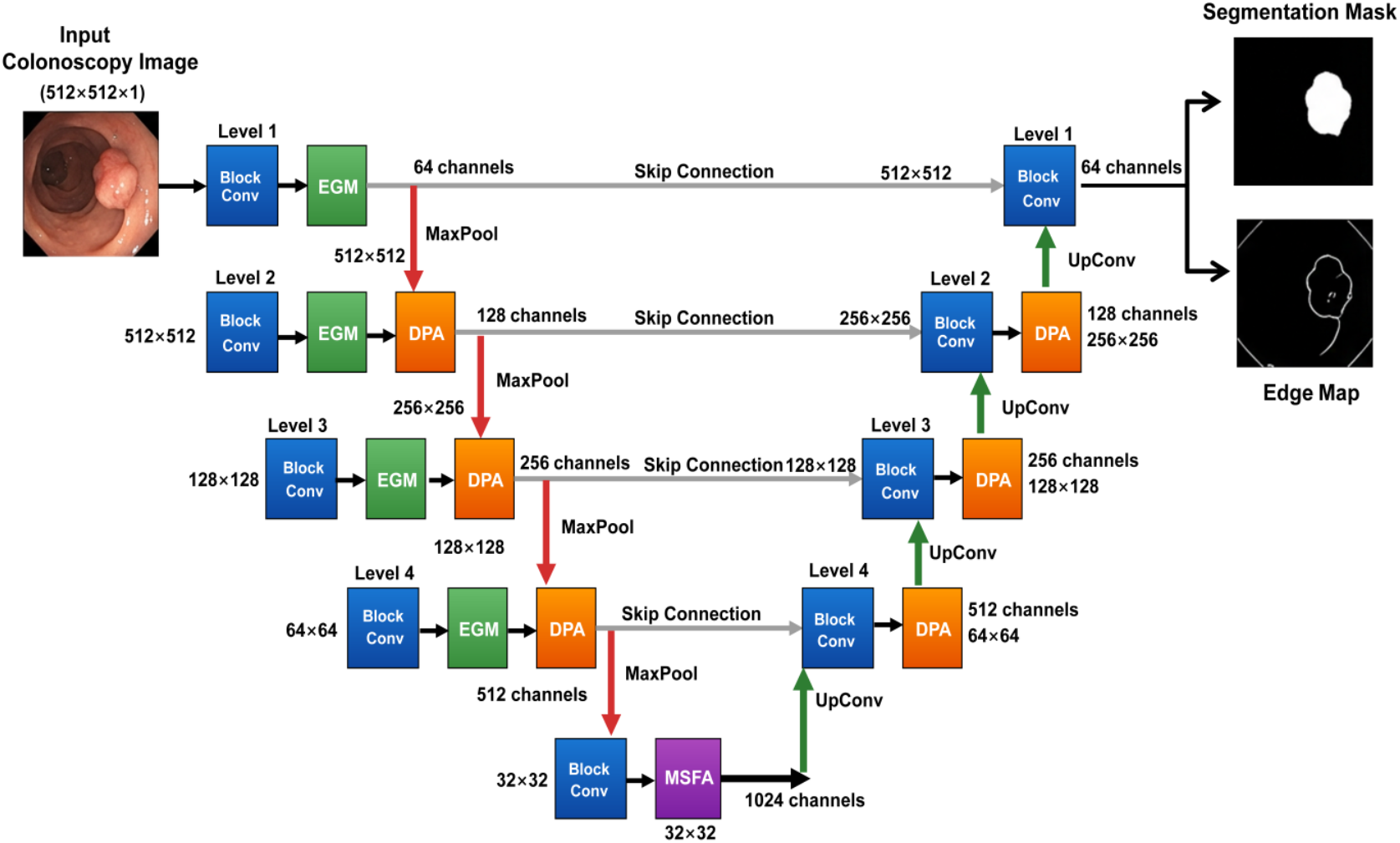
Overall architecture of BEGA-UNet. The encoder incorporates EGM modules for explicit edge extraction at each stage. The bottleneck layer is enhanced with MSFA for multi-scale context aggregation. The decoder path includes DPA modules for feature refinement. Skip connections fuse low-level boundary features with high-level semantic information.

Given input colonoscopy image **I** ∈ ℝ^(3×H×W), BEGA-UNet produces segmentation map **S** ∈ ℝ^(1×H×W) and auxiliary edge prediction **E** ∈ ℝ^(1×H×W). The encoder extracts hierarchical features with channel dimensions [64, 128, 256, 512], each followed by max-pooling. The MSFA module processes bottleneck features (1024 channels). The decoder symmetrically upsamples through transposed convolutions, concatenating encoder features via skip connections.

### 3.2 Edge-Guided Module (EGM)

The Edge-Guided Module (EGM) establishes explicit, learnable boundary representations that constrain the network toward anatomically plausible polyp delineation. The structure is shown in **Figure 2**.

**Figure 2.**
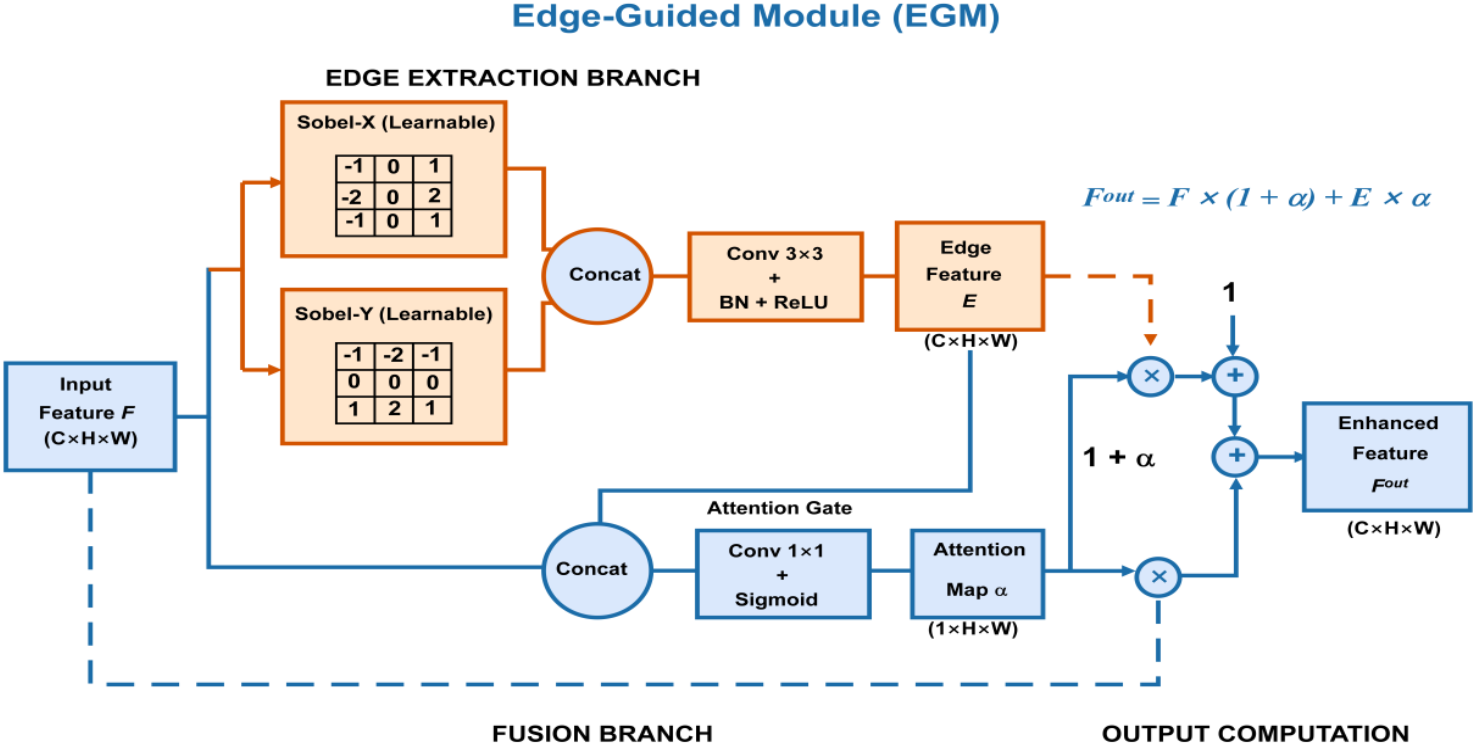
Edge-Guided Module (EGM): learnable Sobel-based operators extract directional edges, fused with semantic features through attention gating.

Given input feature map **F** ∈ ℝ^(C×H×W), EGM applies learnable directional gradient operators:

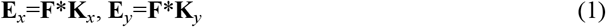

where **K**_*x*_ and **K**_*y*_ are initialized with Sobel kernels but remain learnable. We implement these as depthwise separable convolutions with groups=C, enabling channel-independent edge pattern learning:

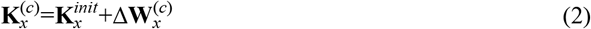

The directional features are fused:

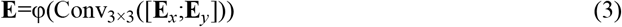

where φ represents Batch Normalization (BN) followed by ReLU.

An attention-based fusion mechanism adaptively integrates edge information:

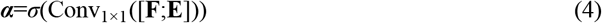

where *σ* denotes the sigmoid activation function.

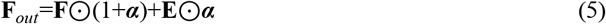

The gating coefficient α adaptively balances semantic and boundary features. The (1+α) formulation ensures that the original feature magnitude is preserved during edge information integration.

### 3.3 Dual-Path Attention (DPA)

Sequential attention mechanisms that apply channel and spatial gating in cascade may attenuate boundary signals as features traverse successive gates. To preserve boundary signal fidelity established by EGM, we propose Dual-Path Attention (DPA), which processes channel and spatial attention in parallel (**Figure 3**), ensuring balanced reinforcement without information bottlenecks.

**Figure 3.**
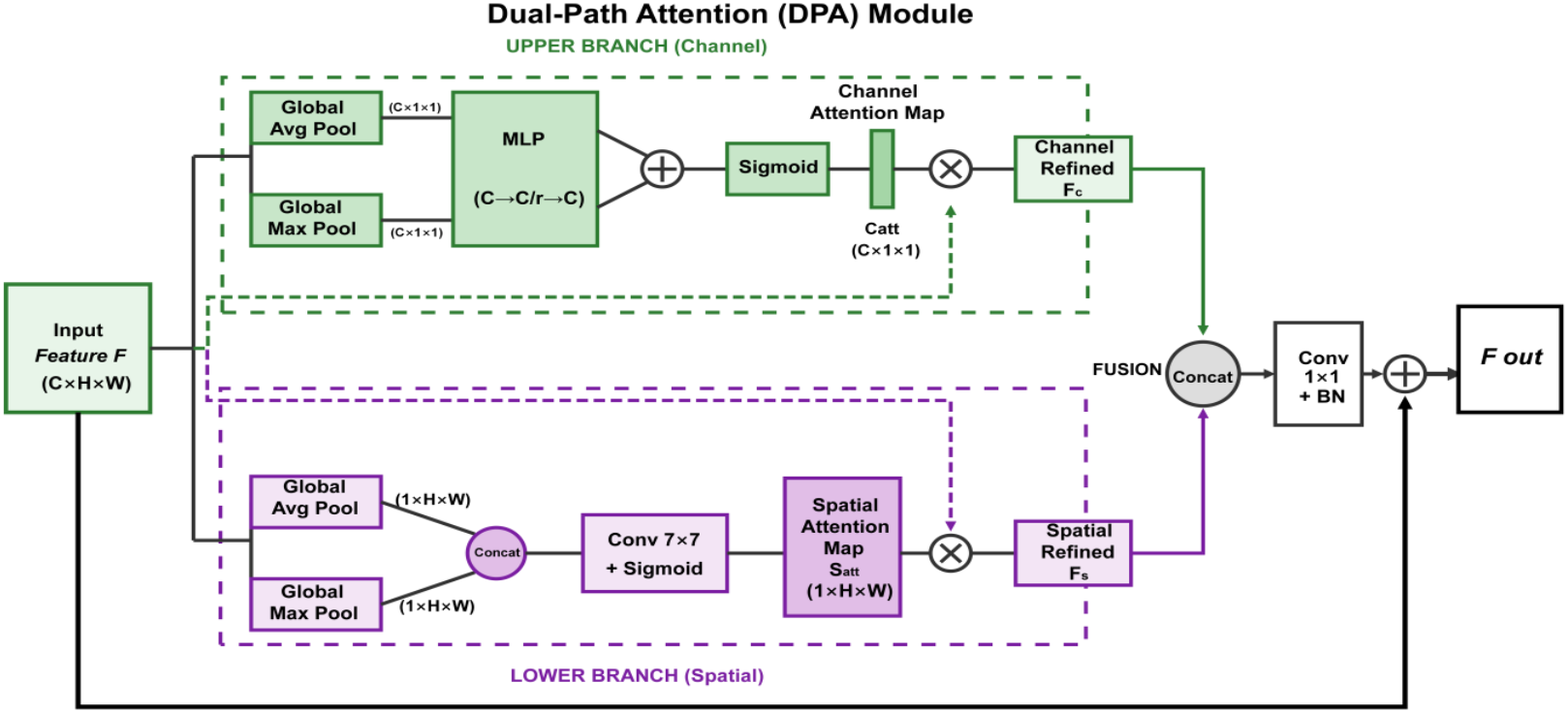
Dual-Path Attention (DPA): channel and spatial attention computed in parallel and fused at feature level.

#### Channel Attention Path

Given **F** ∈ ℝ^(C×H×W), we apply Global Average Pooling (GAP) and Global Max Pooling (GMP) followed by a shared Multi-Layer Perceptron (MLP):

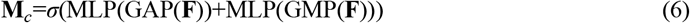

where MLP consists of layers: C → C/16 → C.

#### Spatial Attention Path

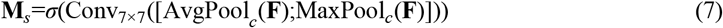

#### Dual-Path Fusion

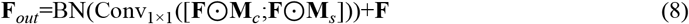

Both paths operate on original features, enabling richer interactions without information loss.

### 3.4 Multi-Scale Feature Aggregation (MSFA)

Dilation rates {1, 2, 4} are empirically selected for 352×352 input resolution(**Figure 4**), differing from DeepLabv3’s {6, 12, 18} designed for higher-resolution natural images. Systematic dilation rate optimization remains for future work.

**Figure 4.**
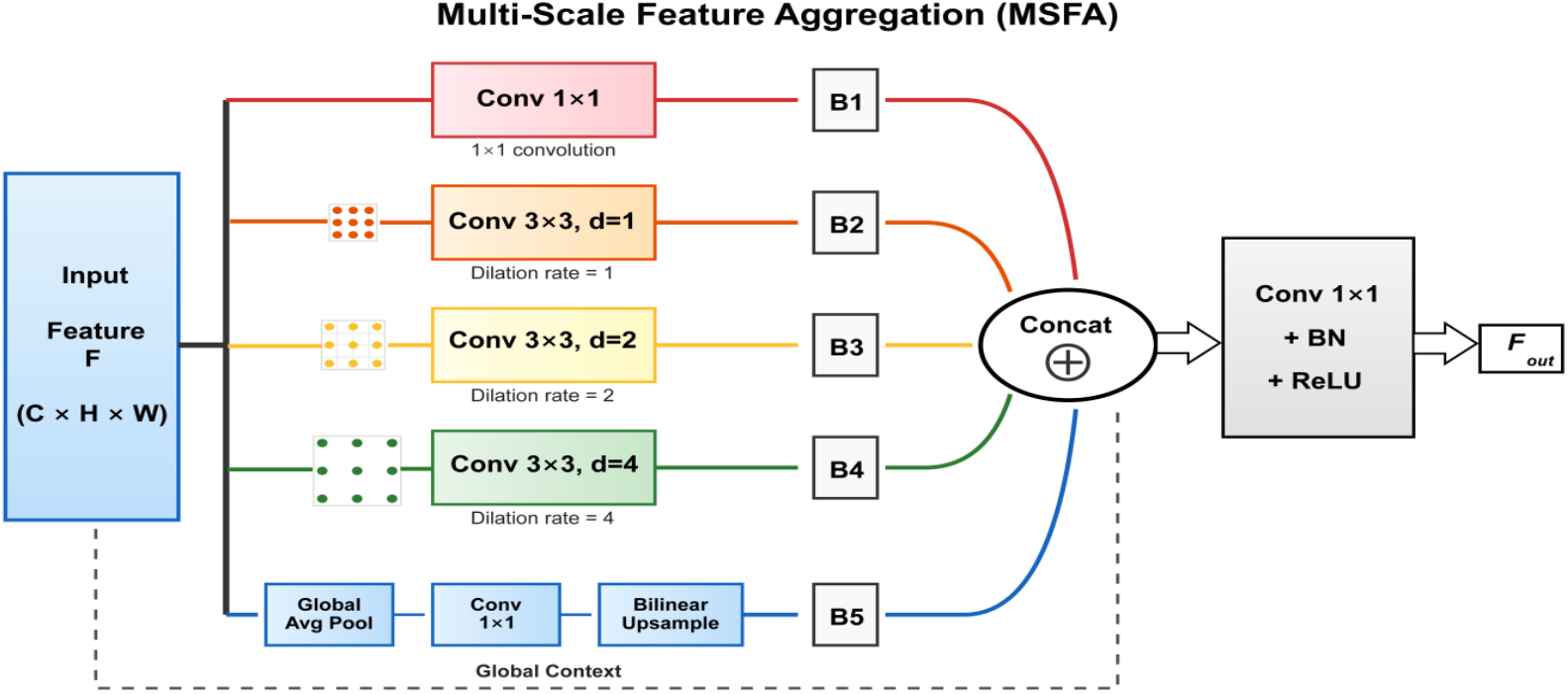
Multi-Scale Feature Aggregation (MSFA): parallel branches with different dilation rates capture multi-scale context.

For input **F** ∈ ℝ^(C×H×W), five parallel branches with varying receptive fields:

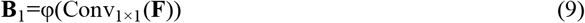

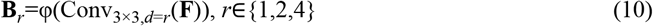

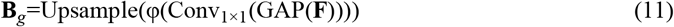

Branches are fused:

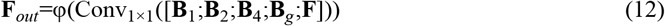

Each branch uses reduced channel dimensions (C/4) to balance representational capacity and computational efficiency.

### 3.5 Loss Function

We adopt a hybrid loss:

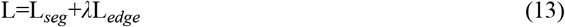

where *Lseg*=*L*_*B*_*CE*+*L*_*D*_*ice* combines Binary Cross-Entropy (BCE) loss and Dice loss, and Ledge = BCE(Epred, B(Y)),where Y denotes the ground truth segmentation mask and B(·) extracts the binary boundary map via morphological gradient. We set λ = 0.2 based on validation performance; sensitivity analysis across λ ∈ {0, 0.05, 0.1, 0.2, 0.3, 0.5} confirms robustness, with Dice varying only 1.74% across the full range (Supplementary Figure S6, Table S4).

## 4. Experiments

### 4.1 Datasets

We evaluate BEGA-UNet on three publicly available polyp segmentation benchmarks. Kvasir-SEG ^[39]^ comprises 1,000 colonoscopy images with pixel-level annotations collected from Vestre Viken Health Trust, Norway. CVC-ClinicDB ^[40]^ contains 612 frames extracted from 29 colonoscopy sequences acquired at the Hospital Clinic of Barcelona. Following established protocols ^[12[22,^ we combine these two datasets (1,612 samples total) and partition them into training (70%, n ≈ 1,128), validation (15%, n ≈ 242), and test (15%, n ≈ 242) sets. Additionally, ETIS-Larib ^[49]^, containing 196 colonoscopy polyp images collected at Larib laboratory, is reserved exclusively for zero-shot evaluation to assess model performance under extreme domain shift scenarios where no training data from the target distribution is available.

Cross-Dataset Evaluation Protocol. To assess generalization under domain shift, we additionally conduct cross-dataset experiments following established protocols ^[45]^. Specifically, we perform bidirectional evaluation: (1) training on full Kvasir-SEG (1,000 images) and testing on full CVC-ClinicDB (612 images), denoted as K→C; and (2) training on full CVC-ClinicDB and testing on full Kvasir-SEG, denoted as C→K. This protocol directly measures model robustness to distribution shift arising from different imaging equipment, patient populations, and clinical centers. We compare BEGA-UNet against three representative baselines: U-Net (classical encoder-decoder), Attention U-Net (attention-enhanced), and TransUNet (Transformer-based), selected to represent distinct architectural paradigms while ensuring training feasibility under identical from-scratch protocols. Methods requiring specialized pre-training (Polyp-PVT, M^2^SNet) or task-specific auxiliary losses (PraNet) were excluded to ensure controlled comparison conditions where architectural contributions can be isolated. All models are trained with identical strong augmentation including aggressive color jittering (brightness, contrast, and saturation variations up to 30%, hue variation up to 10%) to minimize appearance-specific overfitting. Therefore, the observed generalization differences are unlikely to arise from augmentation discrepancies.

### 4.2 Implementation Details

Implementation uses PyTorch 2.0 on NVIDIA RTX 4060 (8GB). Input images are resized to 352×352 pixels and normalized to [0,1]. We employ the AdamW optimizer ^[41]^ with initial learning rate 1×10^−4^, weight decay 1×10^−5^, and cosine annealing schedule over 100 epochs. Batch size is 8 with mixed-precision (FP16) training. Data augmentation includes horizontal/vertical flipping (p=0.5), 90° rotation (p=0.5), affine transformations, Gaussian noise, blur, and color jittering, implemented via Albumentations ^[42]^. Training dynamics (Supplementary Figure S10) demonstrate stable convergence with best validation Dice of 89.37% at epoch 97.

We adopt five evaluation metrics: Dice Similarity Coefficient, Intersection over Union (IoU), Precision, Recall, and 95th percentile Hausdorff Distance (HD95, in pixels). Statistical significance is assessed using Wilcoxon signed-rank tests with Bonferroni correction for multiple comparisons. Effect sizes are quantified using Cohen’s d (thresholds: small d < 0.5, medium 0.5 ≤ d < 0.8, large d ≥ 0.8), and 95% confidence intervals are estimated via bootstrap resampling (n = 10,000 iterations).

We compare against thirteen methods spanning three architectural paradigms: **CNN-based methods** (U-Net ^[9]^, SegNet ^[47]^, PSPNet ^[37]^, ResUNet ^[43]^, and MultiResUNet ^[48]^). **Attention-based methods** (Attention U-Net ^[10]^, PraNet ^[12]^, SANet ^[20]^, UACANet ^[23]^, and CaraNet ^[30]^). **Transformer-based methods** (TransUNet ^[13]^, Polyp-PVT ^[22]^, and M^2^SNet ^[46]^).All methods are trained from scratch using identical experimental settings—including data augmentation, optimizer (AdamW, lr=1×10^−4^), and training epochs (100)—to isolate architectural contributions. This protocol may underestimate methods designed with pre-training (TransUNet, Polyp-PVT); consequently, we emphasize cross-dataset generalization as the primary evaluation criterion, with in-distribution benchmarks serving as secondary reference.

## 5. Results and Analysis

### 5.1 Comparison with State-of-the-Art Methods

Table 1 presents quantitative comparisons across thirteen methods spanning CNN, attention-based, and Transformer paradigms. BEGA-UNet achieves the highest Dice (88.53%), IoU (82.51%), Recall (90.29%), and HD95 (28.20 mm), the second-highest Precision (90.35%, marginally below Polyp-PVT’s 90.54%), and yields statistically significant improvements over baselines with effect sizes varying by baseline, as detailed in later subsections.

**Table 1.** Quantitative comparison on combined Kvasir-SEG and CVC-ClinicDB test set (n = 242). Bold: best; underline: second best. HD95 in pixels (lower is better). All methods trained from scratch under identical protocols for fair comparison.

| Method | Params(M) | Dice(%) | IoU(%) | Precision(%) | Recall(%) | HD95 ↓ |
| --- | --- | --- | --- | --- | --- | --- |
| U-Net | 31.04 | 82.38 | 74.37 | 86.27 | 85.77 | 47.01 |
| PSPNet | 28.3 | 86.46 | 79.81 | 88.30 | 88.16 | 33.94 |
| SegNet | 29.5 | 85.06 | 77.79 | 87.98 | 87.19 | 39.47 |
| Att-UNet | 31.4 | 83.95 | 75.81 | 87.87 | 85.51 | 46.66 |
| ResUNet | 32.47 | 79.19 | 69.54 | 83.31 | 81.79 | 62.76 |
| MultiResUNet | 7.2 | 84.25 | 76.70 | 87.42 | 86.08 | 43.84 |
| PraNet | 10.6 | 84.45 | 77.53 | 87.27 | 86.12 | 34.15 |
| SANet | 18.9 | 84.88 | 77.94 | 85.69 | 88.07 | 38.54 |
| TransUNet | 42.15 | 83.91 | 76.10 | 85.58 | 87.03 | 44.10 |
| UACANet | 15.0 | 83.33 | 76.19 | 86.94 | 85.38 | 36.80 |
| CaraNet | 11.1 | 83.16 | 75.31 | 85.43 | 84.95 | 38.74 |
| Polyp-PVT | 5.9 | 86.38 | 80.03 | **90.54** | 86.90 | 31.45 |
| M<sup>2</sup>SNet | 27.8 | 86.03 | 79.91 | 88.25 | 87.31 | 31.83 |
| **BEGA-UNet (Ours)** | 48.41 | **88.53** | **82.51** | 90.35 | **90.29** | **28.20** |

In-Distribution Performance. Table 1 summarizes results on the combined test set. BEGA-UNet achieves the highest Dice (88.53%) and IoU (82.51%) among compared methods. Effect size analysis reveals heterogeneous practical significance: small effects (d = 0.31-0.48) against classical architectures and negligible effects (d = 0.11-0.13) against recent methods, motivating our emphasis on cross-dataset generalization (Section 5.3) where performance differences become more pronounced.

BEGA-UNet (48.41M parameters) is deliberately designed as a capacity-sufficient analysis model for investigating explicit boundary priors, rather than a deployment-optimized architecture. Despite higher parameter count than lightweight alternatives (Polyp-PVT: 5.9M; M^2^SNet: 27.8M), it achieves real-time inference (54 FPS). Thus, the contribution of this work lies in examining how explicit structural priors relate to domain-robust segmentation, rather than proposing a deployment-ready lightweight model. Knowledge distillation for resource-constrained deployment remains a future direction.

The HD95 metric has direct clinical implications for polyp size estimation, which determines surveillance intervals ^[44]^. BEGA-UNet achieves HD95 of 28.20 pixels, representing ∼11% error reduction over the next-best methods (Polyp-PVT: 31.45; M^2^SNet: 31.83). Under typical colonoscopic resolutions, this may correspond to sub-millimeter boundary localization differences, which could be potentially relevant near the 10mm surveillance threshold ^[44]^.

Table 2 presents the significance test results for Dice scores.

**Table 2.** Statistical Significance and Effect Size Analysis for Dice Score Comparisons with State-of-the-Art Methods. [Δ = BEGA-UNet Dice − Method Dice].

| Method | Dice $\Delta$ | P-value | Sig. | Cohen's d | Effect |
| --- | --- | --- | --- | --- | --- |
| ResU-Net | +9.34% | 2.17e-26 | *** | 0.48 | Small |
| U-Net | +6.15% | 1.12e-14 | *** | 0.31 | Small |
| CaraNet | +5.37% | 1.61e-16 | *** | 0.27 | Small |
| UACANet | +5.20% | 5.96e-11 | *** | 0.25 | Small |
| TransUNet | +4.62% | 7.47e-12 | *** | 0.25 | Small |
| Att-UNet | +4.58% | 6.73e-12 | *** | 0.25 | Small |
| MultiResUNet | +4.28% | 2.09e-10 | *** | 0.22 | Small |
| PraNet | +4.08% | 2.07e-11 | *** | 0.20 | Small |
| SANet | +3.65% | 7.33e-09 | *** | 0.19 | Negligible |
| SegNet | +3.47% | 2.14e-10 | *** | 0.19 | Negligible |
| M<sup>2</sup>SNet | +2.50% | 3.74e-03 | ** | 0.13 | Negligible |
| Polyp-PVT | +2.15% | 2.16e-04 | *** | 0.11 | Negligible |
| PSPNet | +2.07% | 6.22e-06 | *** | 0.11 | Negligible |

Notes: Significance levels after Bonferroni correction: \*\*\* $p < 0.001$ , \*\* $p < 0.01$ , \* $p < 0.05$ . Effect size thresholds: negligible ( $d < 0.2$ ), small ( $0.2 \leq d < 0.5$ ), medium ( $0.5 \leq d < 0.8$ ), large ( $d \geq 0.8$ ).

Table 2 summarizes statistical comparisons with state-of-the-art methods. Effect sizes ranged from small (d = 0.31–0.48 vs. classical architectures) to negligible (d = 0.11–0.13 vs. recent methods), with an average Cohen’s d of 0.23. This indicates that while BEGA-UNet reliably outperforms comparison methods statistically, the marginal gains reflect benchmark saturation rather than architectural limitation. Given the saturation of in-distribution benchmarks—where top-performing methods converge within a narrow performance band—we emphasize effect sizes and cross-domain robustness rather than p-values alone as the primary basis for evaluating architectural contributions. The substantial cross-domain performance disparities reported in Section 5.3 provide more discriminative evidence of genuine architectural advantages. Statistical analysis for ablation configurations is integrated with the dual-protocol ablation study in Section 5.2, Table 3.

**Table 3.**
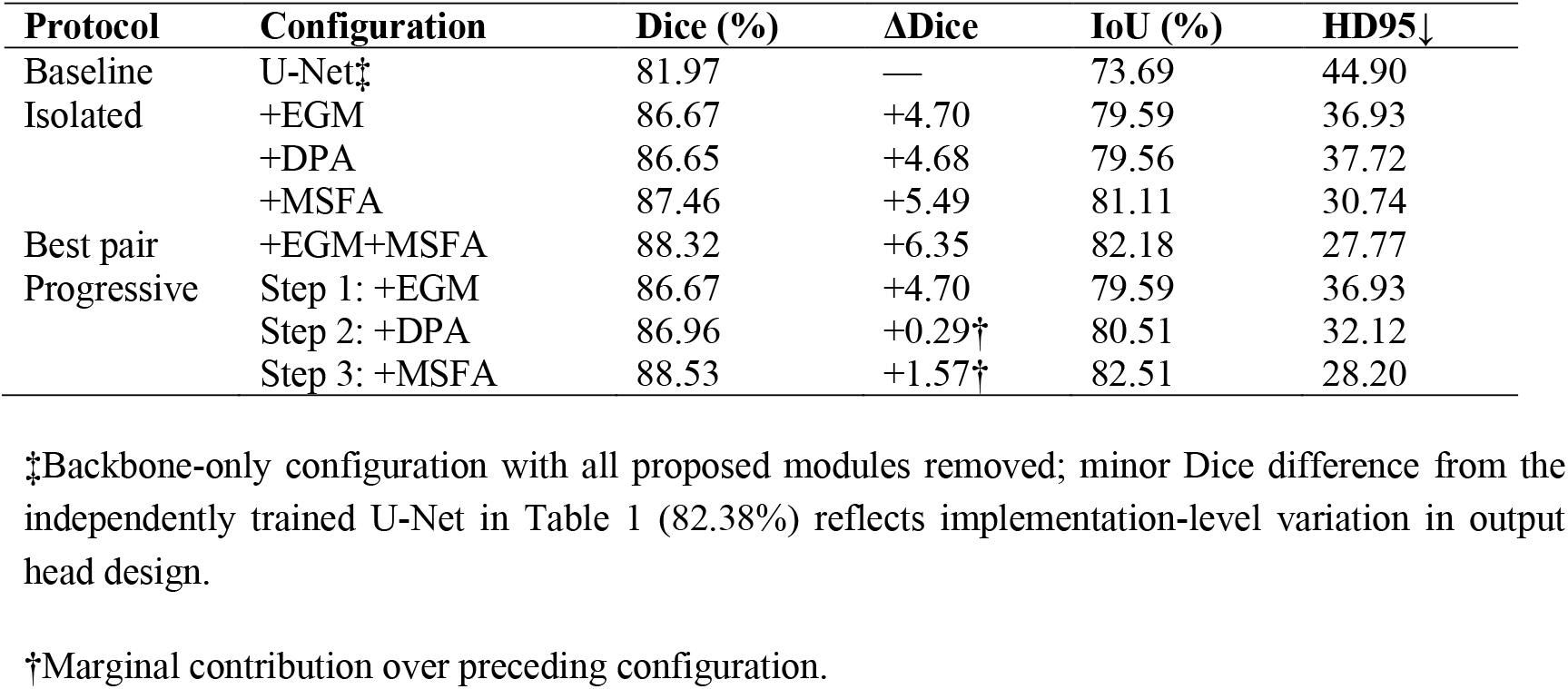
Dual-protocol ablation analysis. Upper: isolated additions to baseline. Lower: progressive integration with marginal contributions. Additivity retention measures realized vs. expected additive improvement for pairwise combinations.

Figure 5 presents multi-faceted performance analysis, including ablation contributions, cross-method comparison, multi-metric radar chart, and parameter-performance trade-off. BEGA-UNet achieves Dice 88.53 ± 4.38% (95% CI: 86.22–90.60%) with consistent improvements across all evaluation dimensions.

**Figure 5.**
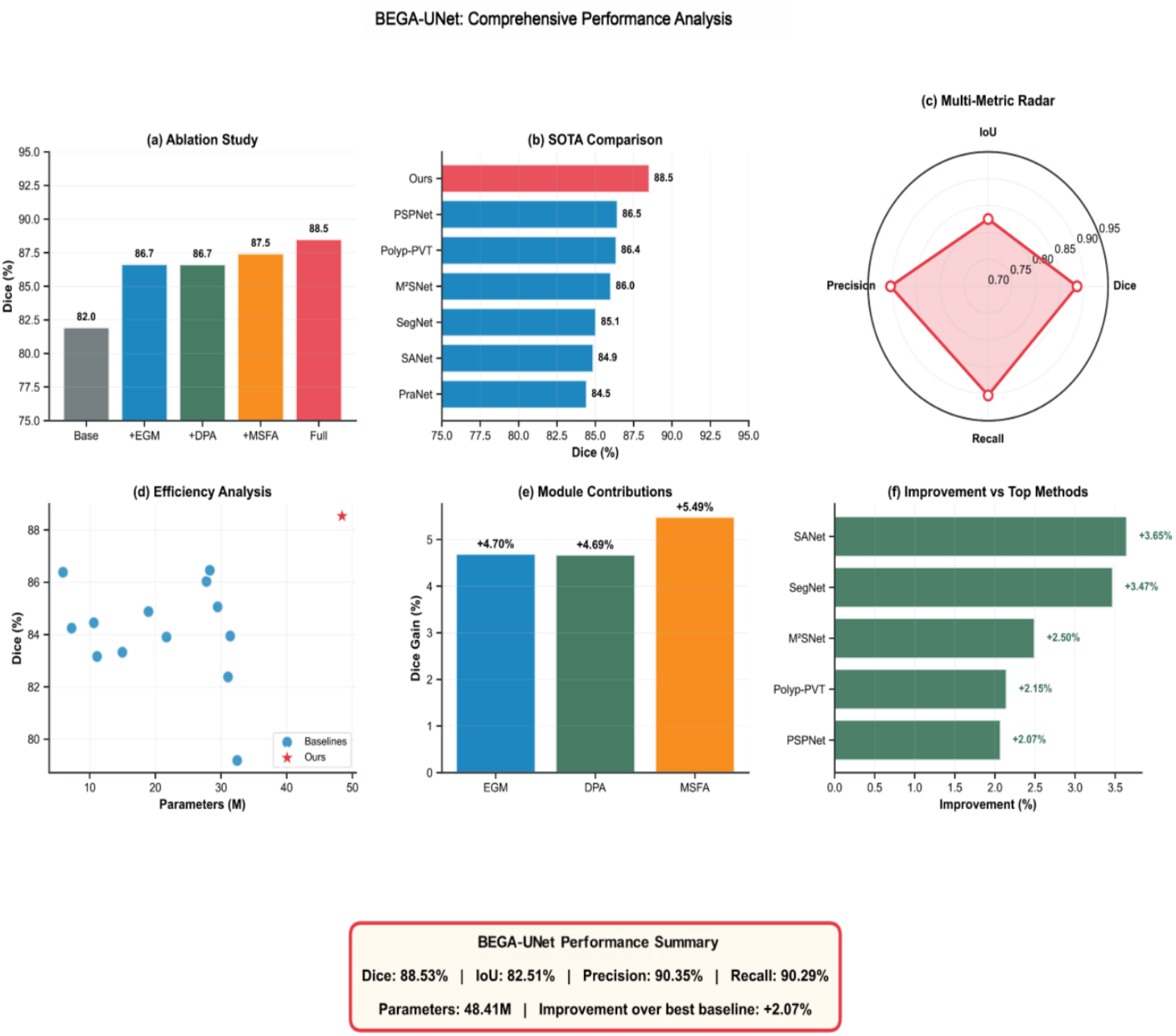
Multi-faceted performance analysis. (a) Ablation study showing module contributions with diminishing returns for full combination. (b) Dice comparison with established methods. (c) Multi-metric radar chart. (d) Parameter-performance trade-off. (e) Individual module contributions. (f) Performance gains over baselines. Summary: BEGA-UNet achieves Dice 88.53±4.38% (95% CI: 86.22-90.60%).

Wilcoxon signed-rank tests with Bonferroni correction confirm statistically significant improvements across all metrics and all 13 baselines (p < 0.05 for all; detailed heatmap in Supplementary Figure S12). Bootstrap confidence intervals (**Figure 6**) further demonstrate stable estimates, with BEGA-UNet’s 95% CI for Dice [0.8622, 0.9060] non-overlapping with most baselines.

**Figure 6.**
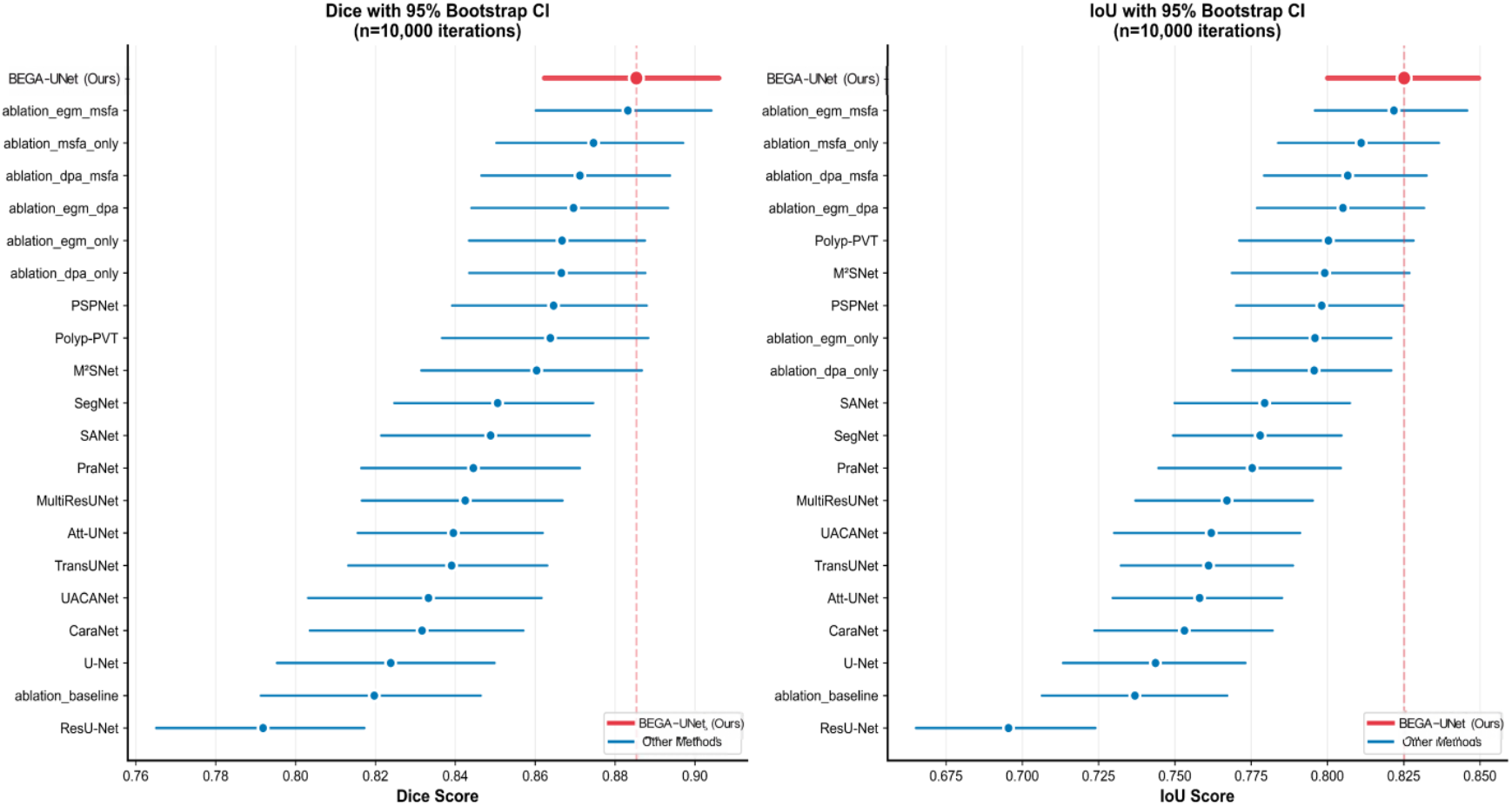
Bootstrap 95% confidence intervals for (a) Dice and (b) IoU scores across all methods (n=10,000 iterations). Red markers and thick lines indicate BEGA-UNet; blue indicates baseline methods. The vertical dashed line marks BEGA-UNet performance. Non-overlapping confidence intervals indicate statistically significant differences.

### 5.2 Ablation Study

We conduct ablation through two complementary protocols—isolated and progressive—to characterize individual contributions and, critically, inter-module functional relationships (**Table 3**). Complete metrics are provided in Supplementary Table S3.

Isolated contributions. Measured against the backbone-only baseline (81.97% Dice; see Table 3 footnote), all three modules yield substantial gains in isolation: MSFA (+5.49%), EGM (+4.70%), and DPA (+4.68%), confirming that each addresses a genuine baseline limitation.

Inter-module interaction analysis. Progressive integration reveals a strikingly different picture: DPA’s marginal contribution drops from +4.68% (isolated) to +0.29% after EGM—a 94% marginal utility reduction. This discrepancy represents a central finding rather than an experimental artifact, which we term functional subsumption: EGM and DPA address the same underlying challenge—boundary signal preservation—through different mechanisms (explicit gradient operators vs. attention-based feature selection). DPA provides substantial gains in the absence of explicit boundary priors, but its marginal utility diminishes to statistical non-significance once boundary constraints are explicitly enforced by EGM.

To quantify pairwise functional overlap, we compute the additivity retention rate *R*=Δ_*p*_*air*/(Δ_*A*_+Δ_*B*_), where Δ_*p*_*air* denotes the actual Dice improvement of the module pair over the baseline, and Δ_*A*_, Δ_*B*_ represent the isolated Dice gains of each single module. Results are summarized in **Table 4**.

**Table 4.** Pairwise module interaction analysis: additivity retention rate and functional relationship characterization.

| Pair | Expected Additive | Actual $\Delta$ | Retention R | Relationship |
| --- | --- | --- | --- | --- |
| EGM+MSFA | 10.19% | 6.35% | 62.3% | Most complementary |
| EGM+DPA | 9.38% | 4.99% | 53.2% | Partial subsumption |
| DPA+MSFA | 10.17% | 5.15% | 50.6% | Strongest subsumption |

EGM+MSFA exhibits the highest additivity retention (62.3%), consistent with their non-overlapping activation patterns (IoU ≈ 0.15–0.20, **Figure 7**): EGM concentrates on boundary pixels while MSFA emphasizes polyp interiors. This functional separation explains why EGM+MSFA achieves 99.8% of full model performance (88.32% vs. 88.53%), with DPA’s residual contribution (+0.21%, p = 0.533) statistically non-significant—suggesting partial functional redundancy rather than architectural deficiency.

**Figure 7.**
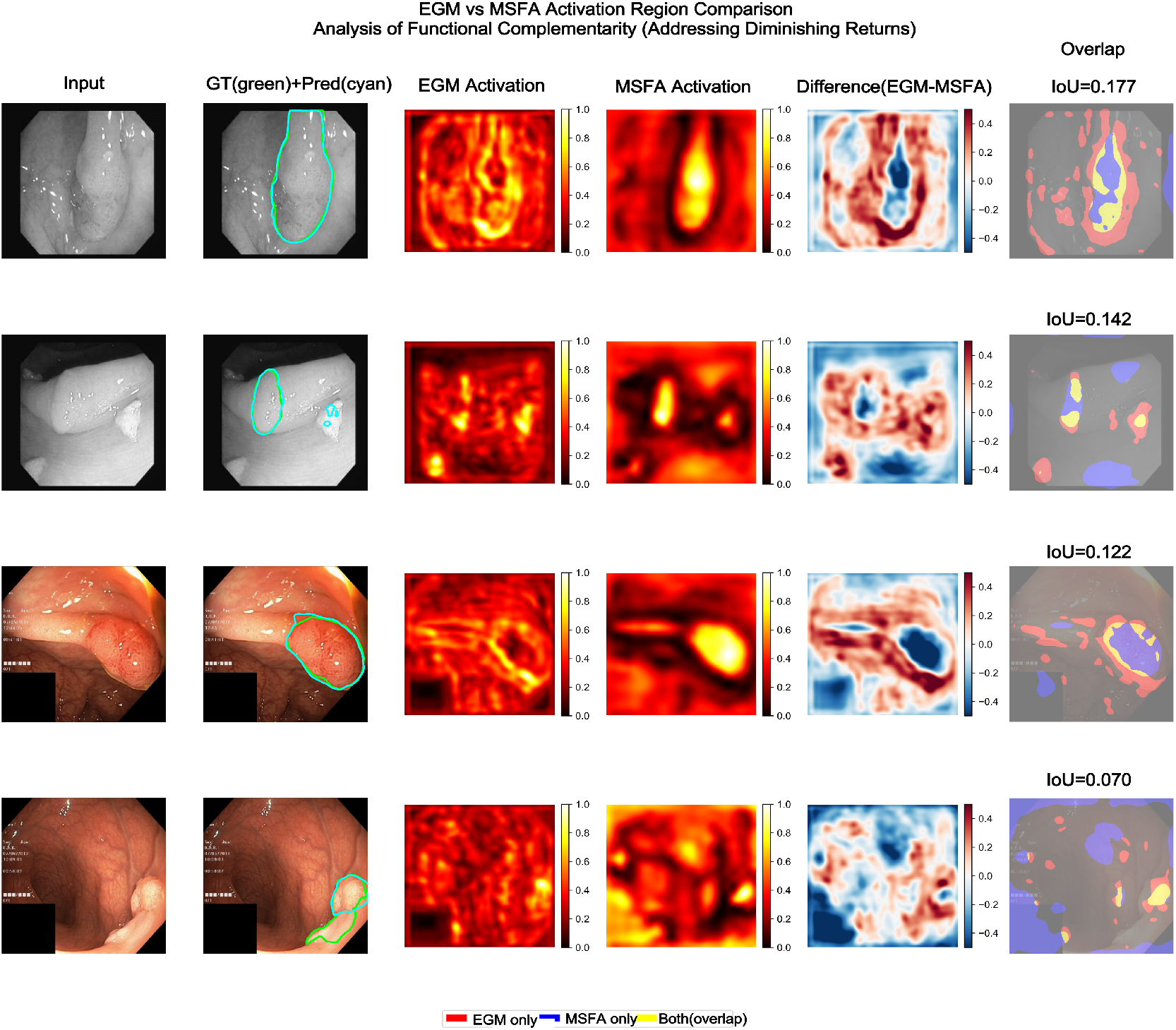
Functional complementarity of EGM and MSFA modules. Columns: input image, GT/prediction overlay, EGM activation (boundary-focused), MSFA activation (interior-focused), difference map (red = EGM-specific, blue = MSFA-specific), overlap visualization with IoU. The distinct activation patterns (∼15% overlap) demonstrate complementary rather than redundant functionality.

Design implications: (1) Explicit boundary modeling functionally subsumes attention-based boundary preservation, favoring dedicated edge operators for boundary-sensitive tasks. (2) Complementary modules should target orthogonal challenges: spatial precision (EGM) vs. scale robustness (MSFA). (3) For resource-constrained deployment, EGM+MSFA provides a principled efficient configuration achieving 99.8% of full model performance.

Having established the individual and combined contributions of each module, we next evaluate the critical question of cross-domain generalization—the primary motivation for explicit boundary modeling.

### 5.3 Cross-Dataset Generalization

Cross-dataset generalization constitutes the primary contribution of this work. Given the saturation of in-distribution benchmarks (Section 5.1), domain shift evaluation provides more discriminative evidence of architectural advantages.

#### 5.3.1 Quantitative Results

Table 5 presents cross-dataset evaluation results. BEGA-UNet demonstrates substantial generalization advantages across both transfer directions.

**Table 5.** Cross-dataset generalization results. K→C: trained on Kvasir-SEG, tested on CVC-ClinicDB; C→K: vice versa. Bold indicates best performance. HD95 in pixels (↓ lower is better).

| Model | Dice_K→C | IoU_K→C | HD95_K→C | Dice_C→K | IoU_C→K | HD95_C→K |
| --- | --- | --- | --- | --- | --- | --- |
| BEGA-UNet (Ours) | 70.33±28.80 | 60.66±29.41 | 41.99±52.15 | 77.04±24.66 | 67.91±26.64 | 34.15±48.49 |
| U-Net | 54.70±31.77 | 44.03±29.69 | 62.63±55.34 | 51.49±29.44 | 39.89±26.71 | 65.01±49.99 |
| Attention U-Net | 44.59±33.55 | 35.12±30.25 | 63.12±49.29 | 35.20±31.83 | 26.49±26.95 | 78.22±55.84 |
| TransUNet | 50.37±29.79 | 39.24±28.49 | 66.50±51.48 | 38.70±27.31 | 27.80±22.79 | 84.31±52.93 |

Table 5 presents bidirectional cross-dataset results. In the K→C direction, BEGA-UNet achieves 70.33% Dice, outperforming U-Net (+15.63%), Attention U-Net (+25.74%), and TransUNet (+19.96%), with 33–37% HD95 error reduction. The C→K direction exhibits even larger disparities: BEGA-UNet achieves 77.04% Dice versus 51.49% (U-Net), 35.20% (Attention U-Net), and 38.70% (TransUNet). Notably, attention-based methods degrade more severely than standard U-Net, suggesting that learned attention patterns may overfit to source-domain appearance characteristics.

#### 5.3.2 Analysis of Generalization Mechanisms

These results are consistent with our central hypothesis: explicit boundary modeling provides domain-invariant structural priors that transfer effectively across imaging conditions. While appearance characteristics (color, texture) vary between datasets due to equipment differences, anatomical boundary geometry—arising from intrinsic tissue properties—remains consistent. EGM’s learnable Sobel operators extract these structural features explicitly, providing representations less susceptible to domain-specific variations. We formalize this observation as the Shape Conservation Hypothesis in Section 6.1.1.

#### 5.3.3 Performance Retention Analysis

Figure 8 visualizes the performance gap between in-distribution and cross-dataset evaluation.

**Figure 8.**
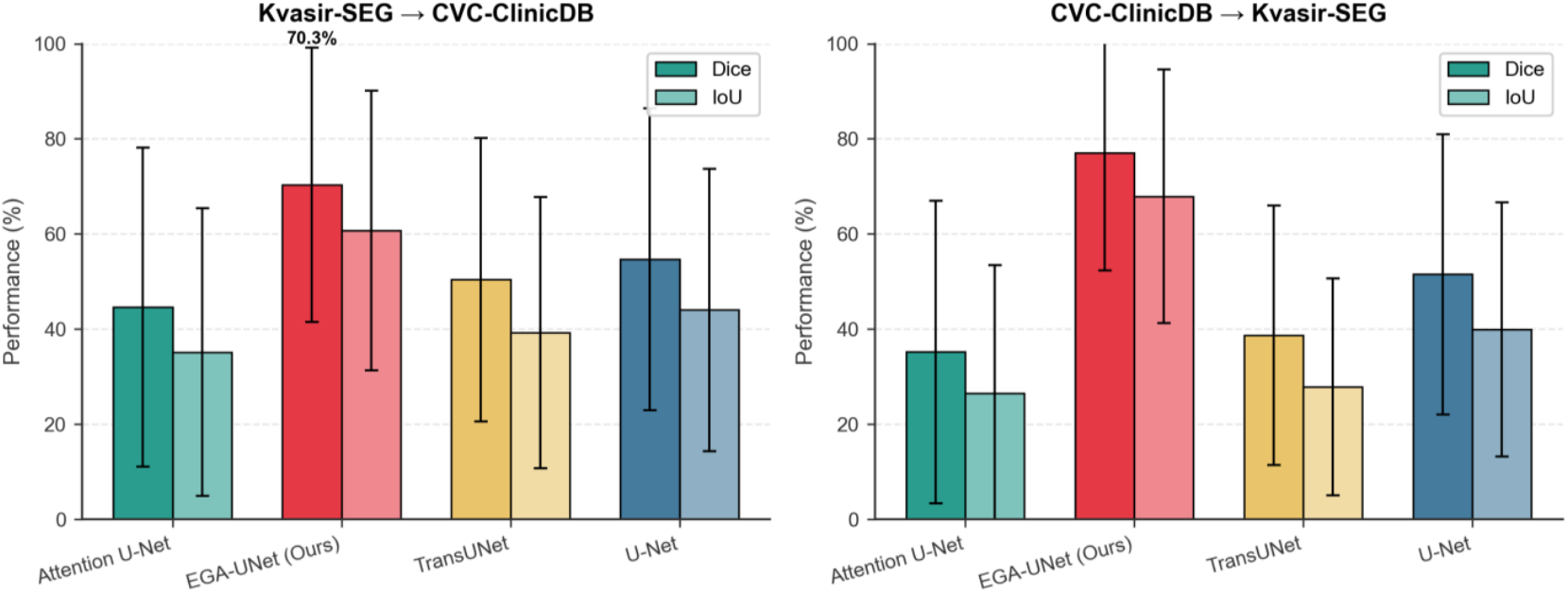
Cross-dataset generalization evaluation. (a) Kvasir-SEG → CVC-ClinicDB transfer. (b) CVC-ClinicDB → Kvasir-SEG transfer. Error bars indicate standard deviation. BEGA-UNet substantially outperforms all baselines in both directions.

BEGA-UNet exhibits the smallest generalization gap: from 88.53% (in-distribution) to 73.69% (cross-dataset average), a drop of 14.84 percentage points. In contrast, U-Net drops from 82.38% to 53.10% (−29.28 points), Attention U-Net from 83.95% to 39.90% (−44.05 points), and TransUNet from 83.91% to 44.54% (−39.37 points). The generalization gap ratio (cross-dataset / in-distribution) quantifies relative robustness: BEGA-UNet maintains 83.2% of its in-distribution performance under domain shift, compared to 64.5% (U-Net), 47.5% (Attention U-Net), and 53.1% (TransUNet). We emphasize that these relative retention ratios—rather than absolute Dice values—constitute the primary evidence for our architectural claims, as they isolate domain robustness from baseline performance level and provide a fairer comparison across methods with heterogeneous in-distribution accuracies.

To assess generalization under more extreme distribution shift, we conducted zero-shot evaluation on ETIS-Larib ^[49]^ (n = 196), a dataset entirely unseen during training originating from different clinical centers with distinct colonoscopic equipment. Without any fine-tuning or domain adaptation, BEGA-UNet achieves 64.3% Dice (median: 81.6%), retaining 72.6% of in-distribution performance. This retention rate—while expectedly lower than the 83.2% observed in cross-dataset experiments—remains substantially higher than baseline methods’ cross-dataset retention (Attention U-Net: 47.5%, TransUNet: 53.1%), indicating that explicit boundary priors maintain discriminative capacity even under severe domain shift. The bimodal performance distribution (Supplementary Figure S13) reflects concentration of failures on flat sessile polyps with minimal boundary contrast, consistent with failure modes identified in Section 5.4.

### 5.4 Qualitative Results and Size-Stratified Analysis

Figure 9 presents representative segmentation results across challenging scenarios, organized by difficulty level from excellent to challenging cases.

**Figure 9.**
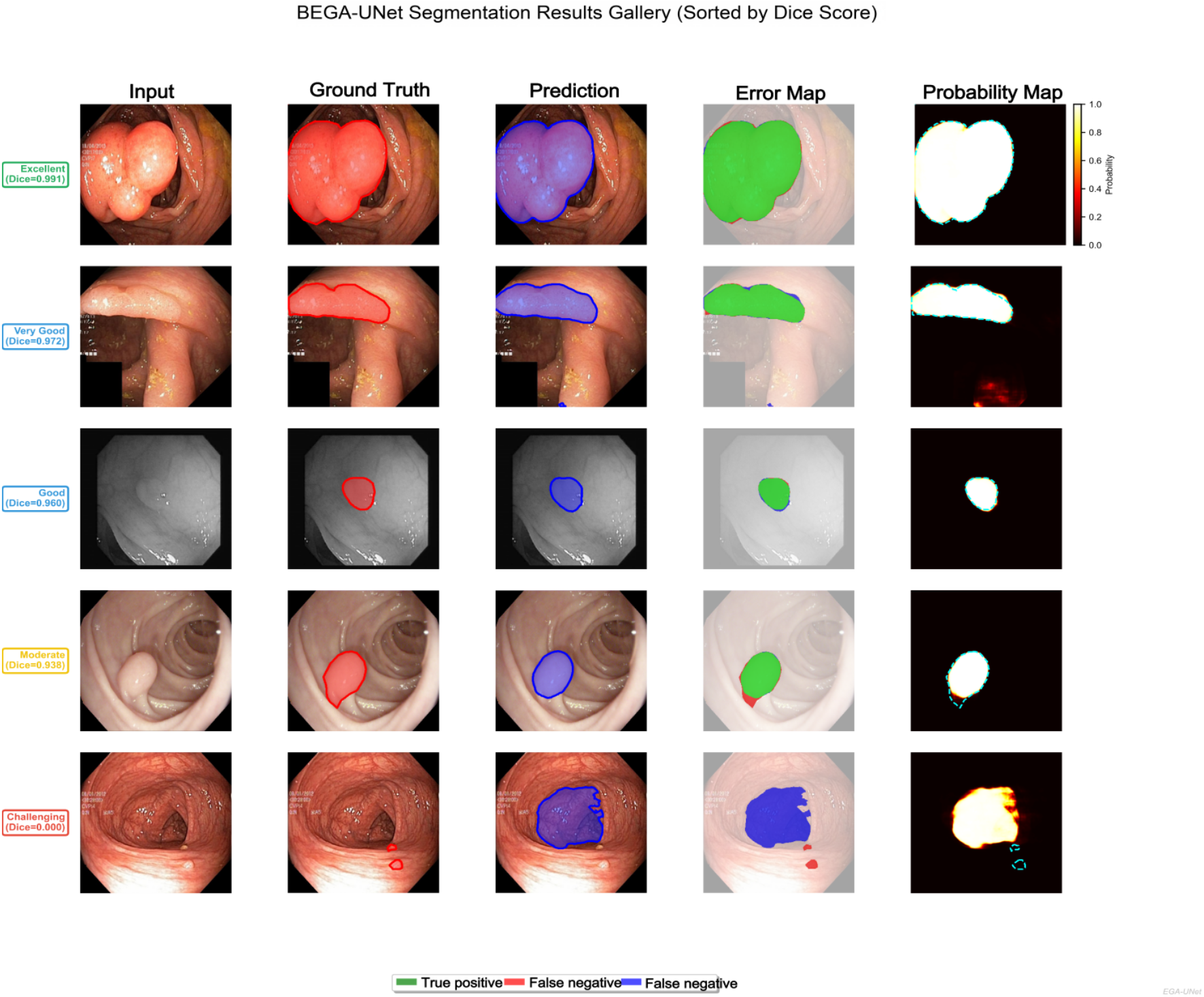
Segmentation Results Gallery (Sorted by Dice Score). Each row presents a case at a different difficulty level (Excellent, Very Good, Good, Fair, Challenging) with corresponding Dice scores; columns from left to right are Input Image, Ground Truth (red annotated polyp region), Prediction (blue predicted polyp region), and Error Analysis (green: True Positive, red: False Negative, blue: False Positive), illustrating the reduction in segmentation accuracy with increasing case difficulty and the corresponding error patterns.

Despite strong overall performance, BEGA-UNet shows limitations for extremely small polyps (<3mm), flat sessile polyps with minimal boundary contrast, and images with severe motion blur (detailed in Supplementary Figure S3). These failure modes highlight opportunities for adaptive boundary modeling in future work.

Size-stratified analysis across the complete dataset (n = 1,612; Supplementary Figure S7) further reveals a clear performance gradient that contextualizes these observations. Large polyps achieve the highest accuracy (Dice: 93.9 ± 8.2%) while small polyps show lower performance (80.6 ± 18.7%) with substantially higher variance, reflecting the geometric challenge that boundary pixels dominate in smaller structures. This size-dependent performance trend aligns with clinical observations of elevated miss rates for diminutive polyps ^[4[5,^ suggesting that the geometric challenges inherent to small polyp segmentation parallel the perceptual challenges faced by endoscopists.

### 5.5 Boundary-Specific Performance Analysis

To quantify boundary delineation precision, we evaluate segmentation accuracy exclusively within narrow bands around the ground truth polyp contour. For each sample, a boundary band *B*_*w*_ is constructed by selecting all pixels whose Euclidean distance to the nearest GT contour point is at most w pixels; Dice and IoU are then computed only within *B*_*w*_. We evaluate band widths w ∈ {2, 5, 8, 10, 15, 20} pixels, corresponding to approximately 0.2–1.7 mm at typical colonoscopic resolution.

Table 6 and **Figure 10** present the results. Several findings emerge. First, all models exhibit monotonic degradation as the evaluation band narrows, reflecting the intrinsic geometric challenge that boundary pixels carry maximal labeling ambiguity. BEGA-UNet’s boundary Dice decreases from 86.93% (w = 20 px) to 62.10% (w = 2 px), while the best-performing baseline (TransUNet) degrades from 83.76% to 60.41% over the same range.

**Table 6.** Boundary-band segmentation performance at varying band widths. Dice (%) reported as mean ± std. Bold: best per band width.

| Method | Overall | w=2px | w=5px | w=10px | w=20px |
| --- | --- | --- | --- | --- | --- |
| BEGA-UNet (Ours) | 88.53 | 62.10±13.44 | 75.42±14.25 | 82.37±14.70 | 86.93±14.71 |
| U-Net | 82.38 | 59.82±16.80 | 71.58±18.14 | 78.03±18.94 | 82.32±19.47 |
| Attention U-Net | 83.95 | 58.76±15.72 | 71.55±16.51 | 78.60±16.81 | 83.35±16.82 |
| TransUNet | 83.91 | 60.41±14.24 | 72.58±15.37 | 79.20±15.91 | 83.76±15.95 |

**Figure 10.**
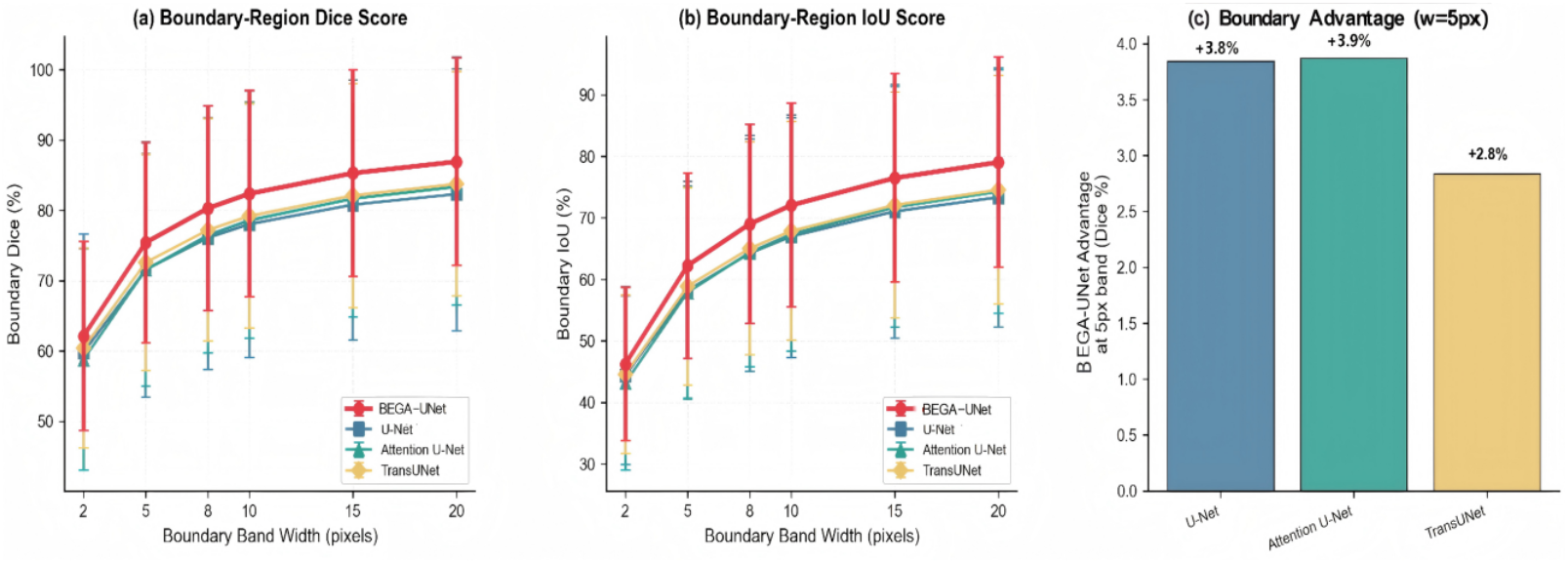
Boundary-band segmentation evaluation. (a) Boundary Dice as a function of band width. (b) Boundary IoU. (c) BEGA-UNet’s absolute Dice advantage over baselines at w = 5 px.

Second, BEGA-UNet achieves the highest boundary accuracy at every band width: +3.84% over U-Net, +3.87% over Attention U-Net, and +2.84% over TransUNet at w = 5 px (Figure 10c). While these margins are more modest than overall Dice differences, they demonstrate consistent boundary-level advantages attributable to explicit edge modeling.

Third, BEGA-UNet’s boundary predictions exhibit lower variance than all baselines. At w = 5 px, BEGA-UNet achieves a coefficient of variation (CV) of 18.9%, compared to 25.3% (U-Net), 23.1% (Attention U-Net), and 21.2% (TransUNet). This consistent reduction in boundary prediction variability indicates that explicit edge modeling produces not only more accurate but also more reliable boundary localization—a property relevant to clinical polyp size estimation, where measurement consistency determines surveillance interval assignment ^[44]^.

### 5.6 Analysis of Edge-Guided Mechanism and Computational Efficiency

Figure 11 visualizes learned edge features at different encoder stages, revealing several key characteristics of the Edge-Guided Module.

**Figure 11.**
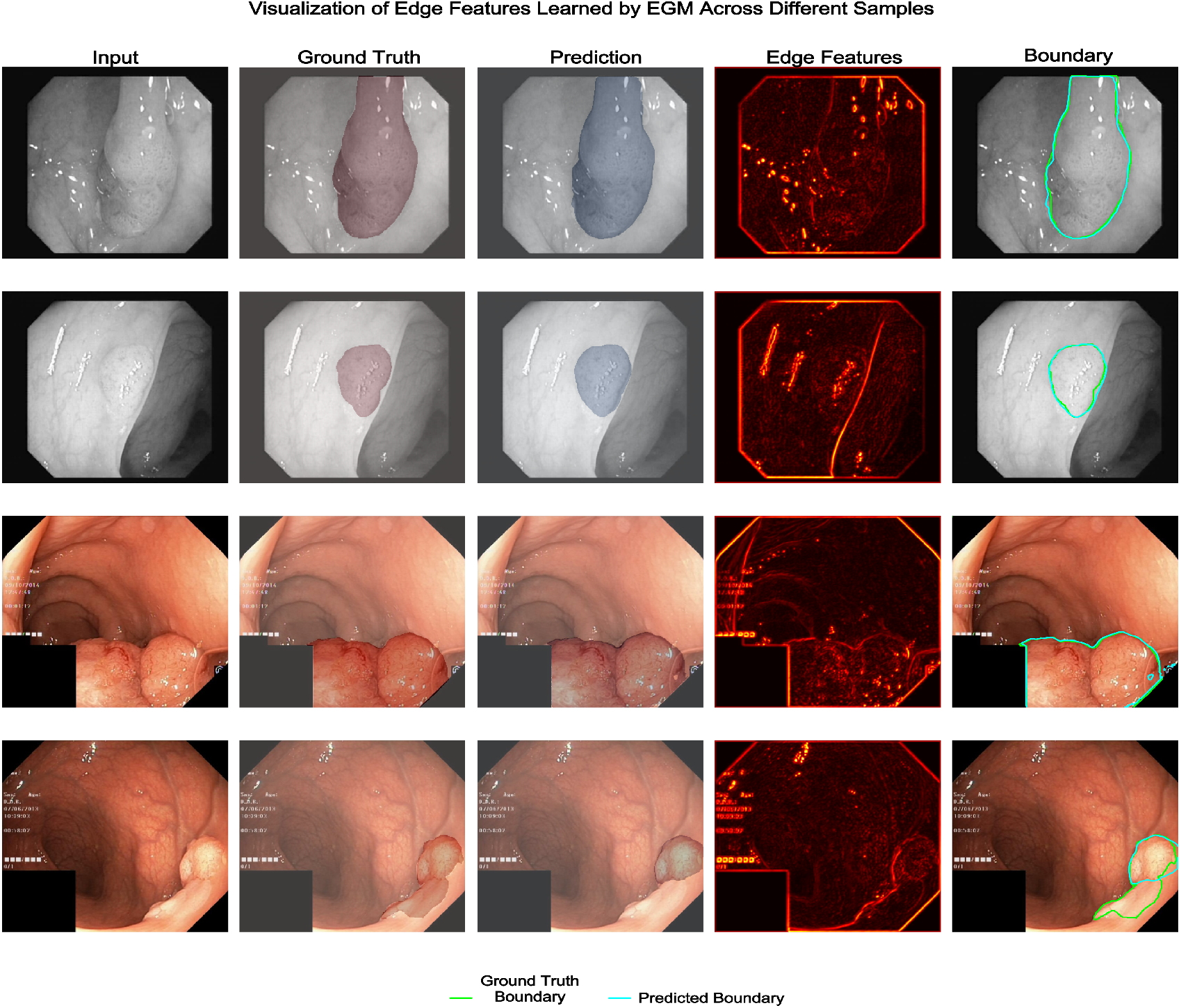
Visualization of edge features learned by EGM across different encoder stages and samples. The module adaptively emphasizes polyp boundaries while suppressing irrelevant edges from vessels and artifacts. Brighter regions indicate stronger edge responses.

Key observations include: (1) Adaptive detection—learned operators selectively emphasize polyp boundaries while suppressing vessel and artifact edges; (2) Hierarchical refinement—shallow layers capture fine textures while deeper layers focus on semantic boundaries; and (3) Task specialization—learned kernels deviate substantially from initial Sobel weights, discovering polyp-specific edge patterns. Supplementary analyses (Figures S4, S8) confirm that EGM adapts to diverse polyp morphologies—both flat and pedunculated—while achieving 85–90% suppression of specular reflection artifacts compared to traditional Sobel operators, validating task-specific structural prior learning.

Regarding computational efficiency, Pareto analysis (Supplementary Figure S9) demonstrates that BEGA-UNet achieves the highest Dice performance among compared methods while maintaining a position on the parameter-performance frontier. As established in Section 5.1, BEGA-UNet is deliberately designed as a capacity-sufficient analysis model rather than a deployment-optimized architecture; despite its 48.41M parameters, it achieves an inference time of approximately 18.5 ms per image (54 FPS), satisfying real-time clinical requirements. The parameter overhead primarily stems from the multi-scale MSFA module essential for handling variable polyp sizes. For resource-constrained deployment, the EGM+MSFA configuration (46.94M, achieving 99.8% of full model performance) offers a principled efficient alternative, and knowledge distillation targeting compact variants remains a priority for future work.

## 6. Discussion

### 6.1 Boundary Features as Domain-Invariant Representations

Cross-dataset experiments (Section 5.3) establish that explicit boundary modeling provides domain-invariant structural priors. We formalize this observation as the Shape Conservation Hypothesis and provide mechanistic validation through feature distribution analysis.

The EGM design instantiates this principle through Sobel-initialized learnable operators. The initialization provides inductive bias toward gradient-based edge detection, while end-to-end learning enables task-specific adaptation. Edge visualizations (Figure 11) demonstrate selective emphasis on polyp boundaries over irrelevant structures (vessels, mucosal folds), suggesting that learned operators discover polyp-specific boundary patterns beyond generic edge detection.

Compared to implicit boundary approaches (M^2^SNet’s multi-scale subtraction), explicit modeling via dedicated operators achieves stronger cross-dataset transfer, though with higher parameter cost. This trade-off warrants further investigation.

#### 6.1.1 Shape Conservation: Mechanistic Explanation

We formalize the Shape Conservation Hypothesis (SCH) to provide a mechanistic explanation for EGM’s cross-domain robustness:**Definition 1 (Shape Conservation Hypothesis)**. Let *D*_*s*_ and *D*_*t*_ denote source and target imaging domains. Across domains, appearance distributions shift significantly:

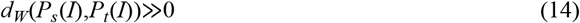

while anatomical boundary geometry remains statistically more stable:

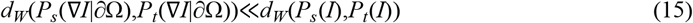

where *d*_*W*_ denotes the Wasserstein distance, ∇I the image gradient field, and ∂Ω the anatomical boundary manifold defined by polyp-mucosa interfaces.

The hypothesis rests on a fundamental asymmetry: endoscopic systems produce systematic color biases from sensor and processing variations, yet edge magnitudes remain approximately invariant under such linear color transformations. More fundamentally, polyp-mucosa boundaries arise from intrinsic tissue properties (vascularization, surface texture, mucosal elevation) determined by pathology rather than imaging equipment.

Figure 12 provides empirical support for SCH: RGB intensity distributions between Kvasir-SEG and CVC-ClinicDB differ substantially (mean per-channel dW ≈ 0.036, ranging from 0.032 to 0.043 across individual color channels), reflecting equipment-specific rendering. In contrast, edge magnitude distributions at polyp boundaries show near-complete overlap (dW ≈ 0.002)—representing a ∼17× reduction in mean cross-domain divergence (up to ∼21× for the most divergent channel), directly satisfying the inequality posited in Definition 1. This quantitative evidence is consistent with our central design principle: EGM’s Sobel-initialized learnable operators extract representations aligned with relatively domain-stable boundary structures, which may help explain why explicit edge modeling exhibits improved robustness compared to purely appearance-driven representations.

**Figure 12.**
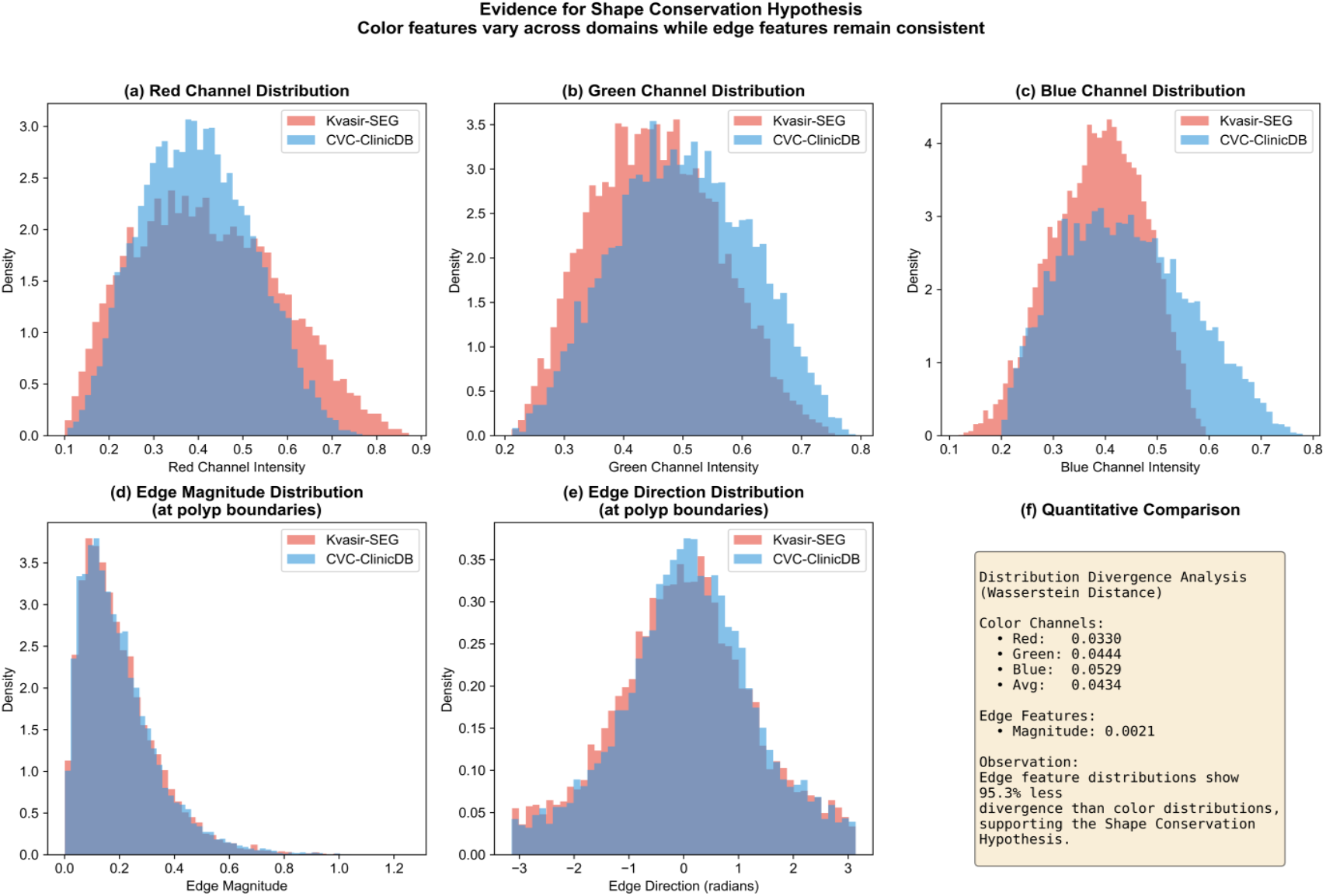
Evidence for Shape Conservation Hypothesis. (a–c) RGB intensity distributions differ between datasets. (d–e) Edge magnitude and direction distributions show near-complete overlap. (f) Quantitative comparison of cross-domain Wasserstein distance: edge magnitude divergence (dW = 0.002) is ∼17× lower than the mean color channel divergence (dW = 0.036). The ∼21× reduction and 95% annotation in panel (f) refer to the comparison against the most divergent individual channel (Blue, dW = 0.043).

To extend this analysis to learned representations, we quantify domain divergence across three feature hierarchies at polyp boundary regions (±5 pixel band): RGB pixels, traditional Sobel gradients, and learned EGM outputs (**Figure 13**). Sobel gradients exhibit minimal domain gap (Maximum Mean Discrepancy, MMD = 0.005), confirming inherent invariance of gradient features. Learned EGM features occupy an intermediate position (MMD = 0.043)—an 11.7× reduction from RGB (MMD = 0.504)—reflecting a principled trade-off: the Sobel initialization provides a domain-invariant foundation, while end-to-end learning adds task-specific discriminability at modest invariance cost. EGM features thus occupy the optimal operating point in the invariance–discriminability trade-off, achieving 11.7× less MMD than appearance features while retaining the boundary selectivity that fixed Sobel operators lack.

**Figure 13.**
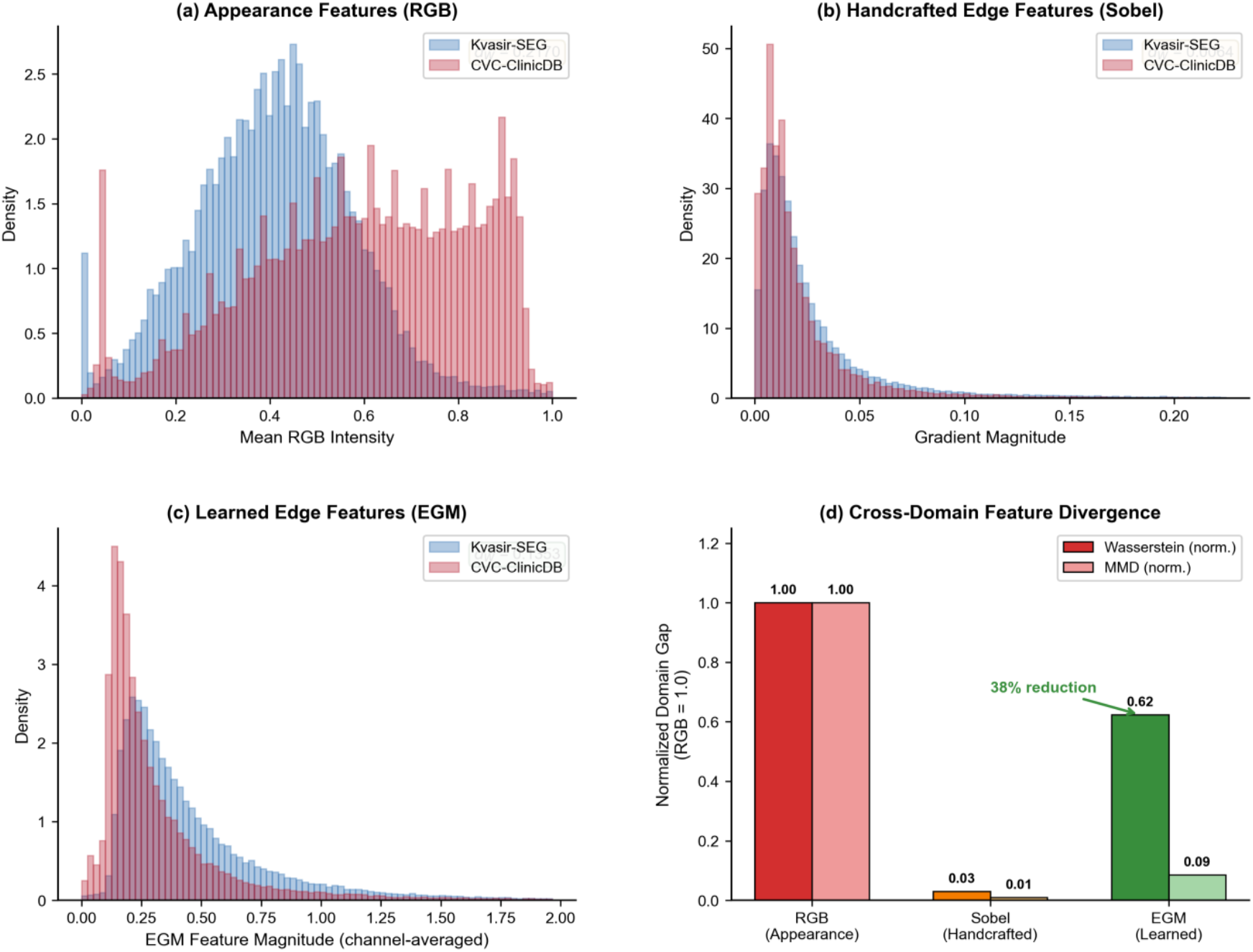
Feature hierarchy domain gap analysis. (a) RGB distributions. (b) Sobel gradient distributions. (c) Learned EGM feature distributions. (d) Normalized domain gap comparison showing 11.7× MMD reduction from RGB to EGM features.

Falsifiability Conditions. The Shape Conservation Hypothesis predicts specific boundary conditions under which explicit edge modeling may lose its domain-invariance advantage: (1) Alternative imaging modalities—under narrow-band imaging (NBI) or chromoendoscopy, artificially enhanced color contrast may create modality-specific gradient patterns that violate cross-domain gradient stability; whether *d*_*W*_(∇*I*|*∂*Ω) remains low under such spectral transformations requires empirical validation. (2) Severe motion artifacts—extreme motion blur destroys high-frequency gradient information entirely, potentially eliminating the structural priors that EGM relies upon; preliminary failure analysis (Section 5.4, Supplementary Figure S3) confirms elevated error rates under motion blur, consistent with this predicted limitation. These boundary conditions delineate a research agenda for adaptive edge modeling that adjusts prior strength based on local image quality assessment.

### 6.2 Architectural Insights and Clinical Implications

The functional subsumption revealed by dual-protocol ablation (Section 5.2) carries broader design implications for boundary-aware architectures. For boundary-sensitive segmentation tasks, our findings suggest that dedicated edge operators should be preferred over attention-based boundary preservation mechanisms. While attention modules provide substantial independent gains, their marginal utility diminishes dramatically (94% reduction) once explicit boundary constraints are enforced through modules like EGM. A complementary finding from cross-dataset experiments is that attention mechanisms (Attention U-Net, TransUNet) exhibit greater vulnerability to domain shift than standard convolutions, with performance drops of 44.05% and 39.37% versus 29.28% for U-Net. This pattern suggests that learned attention weights may overfit to source-domain appearance characteristics, whereas boundary-guided attention mitigates such overfitting by anchoring feature selection to structurally invariant representations.

These architectural findings have potential clinical implications. Improved boundary localization directly affects polyp size measurement accuracy, which in turn informs surveillance interval decisions according to current guidelines ^[44]^. However, we acknowledge that immediate clinical translation faces several barriers: (1) our evaluation remains restricted to European datasets acquired under standard white-light colonoscopy protocols; (2) we have not validated the relationship between segmentation metrics and clinical measurement accuracy; and (3) prospective clinical trials measuring adenoma detection rate impact remain essential. These limitations underscore that benchmark performance, while necessary for methodological validation, is insufficient for clinical adoption.

### 6.3 Limitations and Future Directions

We organize methodological limitations into four categories to contextualize our contributions and guide future research.

#### Evaluation and Generalization

Our cross-dataset and zero-shot validation demonstrates generalization advantages across three benchmarks (Kvasir-SEG, CVC-ClinicDB, ETIS-Larib). However, evaluation remains limited to European datasets with standard white-light colonoscopy. Validation on multi-center datasets (PolypGen), alternative modalities (NBI, chromoendoscopy), and diverse populations constitutes essential future work. The training-from-scratch protocol may underestimate methods designed for transfer learning.

#### Architecture and Translation

BEGA-UNet (48.41M parameters), deliberately designed as a capacity-sufficient analysis model for isolating architectural contributions (Section 5.1), is larger than deployment-optimized alternatives, though real-time inference (54 FPS) meets clinical requirements.Knowledge distillation for embedded deployment and prospective clinical validation measuring adenoma detection rate impact represent essential future directions.Sensitivity analysis across λ ∈ {0, 0.05, 0.1, 0.2, 0.3, 0.5} (Supplementary Figure S6) confirms robustness: Dice varies only 1.74% across the full range (86.87–88.61%), with peak performance at λ = 0.3. The selected λ = 0.2 achieves near-optimal results, with a Dice difference of only 0.08% from the peak, obviating careful tuning for deployment.

#### Future Directions

Four priorities emerge: (1) extended validation on CVC-ColonDB and PolypGen across modalities and diverse populations; (2) knowledge distillation targeting <15M parameters for embedded deployment; (3) temporal modeling exploiting inter-frame consistency for video segmentation; and (4) prospective clinical studies measuring diagnostic concordance and detection rate impact.

Benchmark performance represents an intermediate milestone rather than terminal objective. The pathway from architectural innovation to clinical utility requires addressing each direction, and we hope this systematic analysis—including explicit characterization of module interactions and generalization behavior—provides useful foundation for ongoing efforts.

## 7. Conclusion

We present BEGA-UNet, a polyp segmentation framework grounded in explicit boundary modeling as a structural prior, integrating Edge-Guided Module (EGM), Dual-Path Attention (DPA), and Multi-Scale Feature Aggregation (MSFA). Two principal findings emerge from comprehensive experiments on Kvasir-SEG, CVC-ClinicDB, and ETIS-Larib datasets.

First, explicit boundary modeling substantially enhances domain generalization. BEGA-UNet maintains 83.2% of in-distribution performance under cross-dataset evaluation and 72.6% under zero-shot transfer, substantially exceeding CNN-based (64.5%), attention-based (47.5%), and Transformer-based (53.1%) alternatives. This finding supports the Shape Conservation Hypothesis that boundary features serve as domain-invariant representations. Second, dual-protocol ablation analysis reveals functional subsumption among modules: DPA provides substantial independent gains but undergoes 94% marginal utility reduction once EGM enforces explicit boundary constraints, while EGM+MSFA achieves 99.8% of full model performance, offering a principled configuration for resource-constrained deployment.

In-distribution, BEGA-UNet achieves the highest Dice (88.53%) and IoU (82.51%) among all thirteen compared methods, outperforming the second-best by +2.07% Dice and +2.48% IoU; however, our central conclusions derive from relative performance retention ratios and inter-module interaction patterns rather than absolute benchmark metrics. Future priorities include validation on multi-center datasets across diverse populations and imaging modalities, knowledge distillation for lightweight deployment, and prospective clinical studies correlating segmentation accuracy with adenoma detection rates.

## Acknowledgments

We thank the creators of Kvasir-SEG, CVC-ClinicDB, and ETIS-Larib datasets for making their data publicly available, enabling controlled benchmarking and cross-domain generalization evaluation. We appreciate the open-source implementations of comparison methods that facilitated fair evaluation.

## CRediT Author Statement

Tao Tong: Conceptualization, Methodology, Software, Formal analysis, Investigation, Data curation, Validation, Visualization, Writing – original draft.

Wen Zhang: Validation, Visualization, Writing – review and editing.

Wanni Zu: Supervision, Project administration, Resources, Writing – review and editing.

## Data Availability Statement

The Kvasir-SEG dataset ^[39]^ is publicly available at https://datasets.simula.no/kvasir-seg/. The CVC-ClinicDB dataset ^[40]^ is publicly available at https://polyp.grand-challenge.org/CVCClinicDB/. The ETIS-Larib dataset ^[49]^ is publicly available at https://polyp.grand-challenge.org/EtisLarib/. The source code for BEGA-UNet is available at https://github.com/tongtao-lnu/BEGA-UNet.

## Supplementary Materials

Additional experimental results are provided in the Supplementary Materials document, including extended statistical analyses (Figures S1–S13) and detailed numerical results (Tables S1–S7).

## Declaration of generative AI and AI-assisted technologies in the manuscript preparation process

Statement: During the preparation of this work the authors used Claude (Anthropic) in order to assist with language polishing, text refinement, and improving the clarity of the manuscript. After using this tool, the authors reviewed and edited the content as needed and take full responsibility for the content of the published article.

